# Antibiotic resistance profile of *E. coli* isolates in 17 municipal wastewater utilities across Oregon

**DOI:** 10.1101/2021.11.15.21266366

**Authors:** Marjan Khorshidi-Zadeh, Sue Yee Yiu, Jacquelynn N. Nguyen, Gabriela L. Garza, Joy Waite-Cusic, Tyler S. Radniecki, Tala Navab-Daneshmand

## Abstract

Wastewater treatment utilities are considered one of the main sources and reservoirs of antimicrobial resistance. The objective of this study was to determine the diversity and prevalence of antibiotic-resistant *Escherichia coli* in wastewater treatment systems across the state of Oregon. Influent, secondary effluent, final effluent, and biosolids were collected from 17 wastewater treatment utilities across Oregon during the winter and summer seasons of 2019 and 2020 (*n* = 246). *E. coli* strains were recovered from samples by culturing on mTEC, followed by confirmation with MacConkey with MUG agar plates. Antibiotic susceptibility of 1143 *E. coli* isolates against 8 antibiotics were determined, and resistance profiles and indices were analyzed between utilities, seasons, and flows. Antibiotic resistance phenotypes were detected in 31.6% of the collected *E. coli* isolates. Among those antibiotic-resistant *E. coli* isolates, multi-drug resistance (i.e., resistance to three or more classes of antibiotics) was harbored by 27.7% with some strains showing resistance to up to six classes of antibiotics. The most prevalent resistance was to ampicillin (*n* = 207) and the most common combinations of multi-drug resistance included simultaneous resistances to ampicillin, streptomycin, and tetracycline (*n* = 49), followed by ampicillin, streptomycin, and sulfamethoxazole/trimethoprim (*n* = 46). Significant correlations were observed between resistance to sulfamethoxazole/trimethoprim and resistances to ampicillin, ciprofloxacin, and tetracycline (*p* < 0.001). A small percentage (1.1%) of the *E. coli* isolates displayed extended-spectrum beta lactamase (ESBL) activity and a single isolate carried resistance to imipenem. Compared to wastewater influent, ciprofloxacin resistance was significantly more prevalent in biosolids (*p* <0.05) and tetracycline resistance was significantly lower in effluent (*p* <0.05). Seasonal impact on antibiotic-resistant *E. coli* in wastewater influent was observed through significantly higher multiple antibiotic resistance (MAR) index, ampicillin resistance prevalence, and ciprofloxacin resistance prevalence in summer compared to winter (*p* < 0.05). This state-wide study confirms the widespread distribution of antibiotic-resistant, multi-drug resistant, and extended-spectrum beta lactamase-producing *E. coli* in wastewater systems across different flows and seasonal variations, making them the recipients, reservoirs, and sources of antimicrobial resistance.

## 1. Introduction

Infections with antibiotic-resistant bacteria (ARB) are an urgent and emerging human health concern worldwide due to their association with increased hospital stays, morbidity, and mortality. In the U.S., antibiotic-resistant pathogens are responsible for over 35,000 deaths per year and upwards of $20 billion in direct healthcare costs (CDC, 2019, 2013). Municipal wastewater treatment utilities are hotspots for ARB and their determinant genes (Berendonk et al., 2015). Clinically relevant ARB as well as multi-drug resistance (MDR; resistance to three or more classes of antibiotics) phenotypes are present in municipal wastewater (Hoelle et al., 2019). Of importance are critical priority pathogens defined by the World Health Organization, including extended-spectrum beta-lactamases (ESBL)-producing and carbapenem-resistant Enterobacteriaceae (WHO, 2017). Globally, treated or untreated wastewater and biosolids are used for irrigation and soil amendment in agriculture, making soil and food crops potential conduits for human exposure to pathogenic ARB (Mays et al., 2021). Further, there are reports of presence and dissemination of ARB and their determinant genes in environmental reservoirs, such as rivers, lakes, and sediments associated with wastewater discharge (Logan et al., 2020; Quintela-Baluja et al., 2019). Understanding the parameters that impact ARB survival and persistence in wastewater treatment processes will support the development of evidence-based strategies to combat their negative public health impact.

Different wastewater treatment processes impact microbiological quality and hence the ARB load of final effluent and biosolids differently. The biological wastewater treatment compartment can provide favorable conditions for horizontal and vertical transfer of resistance genes via mobile genetic elements within the same or different taxonomic levels of bacteria (Berendonk et al., 2015). Studies have reported increases in the prevalence of *E. coli* resistant to different antibiotics as well as MDR phenotypes after biological treatments, including activated sludge and lagoons (Kumar et al., 2020; Mezrioui and Baleux, 1994; Zhang et al., 2009). It has been suggested that parameters such as solid retention time, temperature, and type of treatment processes impact ARB levels in biological compartments (Mezrioui and Baleux, 1994; Sui et al., 2017). Disinfection of secondary effluent is suggested to generally reduce ARB levels in the final effluent (Sousa et al., 2017). Subsequently, over 99% of surviving ARB populations in municipal wastewater influent end up in residual biosolids (Burch et al., 2013a). Advanced sludge treatment processes such as lime stabilization, aerobic digestion, and air-drying beds are capable of reducing ARB populations, whereas other processes (e.g., conventional dewatering, gravity thickening, or anaerobic digestion) have shown little impact (Burch et al., 2013b; Munir et al., 2011).

In addition to treatment processes, seasonal variations can impact ARB populations in wastewater influent as well as the final effluent and biosolids. Seasonal changes can be accompanied by considerable temperature variability, precipitation, as well as increased prescription and consumption of antibiotics. Environmental temperature is an important parameter that can impact the overall microbial diversity (Manaia et al., 2018), horizontal gene transfer rates (Miller et al., 2014), and the incidence of antibiotic resistance genes (Sui et al., 2017) in wastewater. Seasonality in antibiotic concentrations has also been reported in the influent received by wastewater treatment utilities (Golovko et al., 2014). Higher rates of antibiotic-resistant *E. coli* in winter (compared to other seasons) have been reported in wastewater influent obtained from two wastewater utilities in France (Mezrioui and Baleux, 1994). Similarly, higher abundance of ESBL-producing *E. coli* have been reported in winter as compared to summer in samples collected from 16 wetlands and three rivers in France (Henriot et al., 2019). Higher precipitation has also been suggested to correlate with increased antimicrobial resistance (AMR) and MDR in enterobacteria in a freshwater watershed in Brazil (Lima-Bittencourt et al., 2007). Others did not observe any significant seasonal associations in ARB populations in an urban-combined sewer system in Japan (Honda et al., 2020). Lack of large-scale studies that include more diverse datasets makes it difficult to arrive at more generalized conclusions from these research findings. There remains a critical need for larger scale, more holistic studies to estimate prevalence and dynamics of ARB in wastewater systems to translate these findings into practical and actionable guidance for the wastewater treatment industry.

The aim of our study was to characterize the antibiotic resistance profile of *E. coli* (as an indicator of enteric bacteria) in wastewater systems across Oregon, and to determine the impacts of seasonal variations and treatment processes on their fate and presence. To achieve these objectives, wastewater influent, secondary effluent, final effluent, and biosolids were collected from 17 wastewater treatment utilities across the state of Oregon in the winter and summer seasons of 2019 and 2020. The AMR phenotypes of 1143 *E. coli* isolates were determined, and resistance profiles and indices were analyzed between utilities, seasons, and flows (i.e., influent, secondary effluent, final effluent, or biosolids).

## 2. Material and methods

### 2.1. Study sites and sample collection

Wastewater treatment utilities (*n* = 17) across the state of Oregon were recruited by invitation in this study (Table S1). Sampling was done over two seasons (summer and winter) in the time span of two years (2019 and 2020). Summer samples were collected between the months of July and September and winter samples were collected between January and March. The enrolled treatment utilities served different population densities ranging between 2,000-80,000 (*n* = 11), and 80,000-700,000 (*n* = 6). The enrolled utilities were located in the coastal region (*n* = 2), Willamette Valley (*n* = 10) or in Eastern Oregon (*n* = 5). The Oregon climate is divided by the Cascade Range into two main regions. The western part of the state (coastal and Willamette Valley) is influenced by the Pacific Ocean, very wet (161.5 mm precipitation) with moderate temperatures (2.7/10.9°C; low/average) in winter and dry (26.6 mm precipitation) with warm temperatures (11.9/24.8°C) in summer (US Climate Data, 2021). The eastern part of Oregon (high desert) has snowy (79.4 mm precipitation), cold (−2.5/8.6 °C) winters and very dry (17.3 mm precipitation) and warm (10.4/28.9 °C) summers (US Climate Data, 2021). The minimum distance between adjacent participating utilities was 8 km and maximum distance was 575 km. Domestic sewage was the main inlet flow to these treatment utilities with the majority receiving less than 20% industrial wastewater (only one utility receives over 80% industrial flow). Of the 17 utilities, 12 receive hospital sewage and 10 receive septic sludge. Finally, only three of the wastewater treatment utilities were combined sewer systems that also receive stormwater. Within the study sites, the average daily flow rates during winter (wet) and summer (dry) seasons ranged from 0.8 to 340.7 and 0.8 to 196.8 million L/day, respectively. Treatment processes in the enrolled utilities included some form of primary treatment followed by secondary treatment. The utilities were categorized into two groups according to the secondary treatment process (Table S1): the majority of utilities (*n* =12) used conventional treatment processes (i.e., lagoon, activated sludge, oxidation ditch), and the rest used an advanced treatment (i.e., biological nutrient removal, membrane bioreactor) (*n* = 5). For disinfection, utilities used either chlorine (*n* = 14) or ultraviolet (UV) treatment (*n* = 4). Treated wastewater effluent is discharged to rivers or the Pacific Ocean (*n* = 13), to wetlands and ponds (*n* = 3), or is used for agricultural irrigation (*n* = 3). Biosolids treatment processes included anaerobic digestion (*n* = 12) as the most common method followed by aerobic treatment (*n* = 2), alkaline treatment (*n* = 2), and mechanical dewatering (*n* = 1). Treated biosolids, if available (*n* = 15), are land applied (*n* = 11), disposed of in landfills (*n* = 2), or both (*n* = 2). Only two of the study sites produce Class A biosolids as defined by the U.S. Environmental Protection Agency (US EPA, 1999). From each wastewater treatment utility, samples of influent (1 L), secondary effluent (after biological treatment and before disinfection; 1 L), final effluent (1 L), and treated biosolids (500 g) were collected. During the four sampling campaigns (winter 2019, summer 2019, winter 2020, and summer 2020) a total of 246 samples were collected from the 17 participating wastewater treatment utilities. Biosolids were not collected from utility F in summer 2019. Additionally, no samples were collected from utility A in winter 2019 nor from utilities A, C, F, and K in summer 2020 (due to challenges associated with the COVID-19 pandemic). Samples were stored on ice and transported or shipped to the laboratory at Oregon State University where they were processed within 24 hours of collection.

### 2.2. Physical and chemical parameters

Total solids, total and volatile suspended solids, pH, and conductivity were measured using standard methods (APHA, 2012; US EPA, 1983). Total solids of biosolids samples were determined by differential mass (starting mass: 2 ± 0.4 g) after drying overnight at 104 ± 1 °C. Total suspended solids were determined for wastewater influent (up to 75 mL) and final effluent (150 mL) by filtering through 1.2 μm glass microfiber filter (Whatman, Kent, UK) followed by drying at 104 ± 1 °C for 2 hours. Volatile suspended solids were determined by differential mass of the filter before and after ignition at 550 °C. Influent and final effluent pH and conductivity were measured (sympHony, Radnor, PA) after mixing on stir plate for 10 min. For biosolids, 2 ± 0.4 g of solids were mixed with 18 mL of deionized water for 20 minutes prior to pH and conductivity measurements. Ammonia levels were measured in wastewater influent and final effluent using Hach method 10031 for high range samples (Hach, Loveland, CO).

### 2.3. E. coli isolation

To isolate *E. coli* colonies from secondary effluent and final effluent, up to 400 mL of well-mixed samples were vacuum filtered on a 0.45 µm mixed-cellulose ester membrane (Whatman, Kent, UK). Filters were placed on m-TEC ChromoSelect agar (Sigma Aldrich, St. Louis, MO) plates. Wastewater influent and biosolids were streaked directly onto m-TEC agar plates. Plates were incubated at 44.5 □ for 22-24 hours. Colonies displaying a purple/magenta color were considered to be presumptive *E. coli*. Typical colonies (1-11 per sample) were transferred to LB broth and incubated at 37 □ for 16-18 hrs. Isolates were streaked onto MacConkey agar with MUG (Hardy Diagnostics, Santa Maria, CA). After incubation at 37 °C for 18-24 hours, isolates were considered to be confirmed as *E. coli* if they were pink in color and exhibited fluorescence under blue-green long-wave UV light (366 nm). Confirmed isolates were cultured in LB broth at 37 °C for 16-18 hours, supplemented with 20% glycerol, and stored at −20 °C for later analysis. The number of isolates collected from each flow (i.e., influent, secondary effluent, final effluent, and biosolids) from each facility during each sampling season are shown in Figure S1.

### 2.4. Antibiotic susceptibility testing

Antibiotic susceptibility phenotypes *of E. coli* isolates (*n* =1143) were determined using the standard disk diffusion technique following the Clinical and Laboratory Standards Institute guidelines (CLSI, 2020). The antibiotics used in this study were ampicillin (10 μg), cefotaxime (30 μg), ceftazidime (30 μg), ciprofloxacin (5 μg), imipenem (10 μg), streptomycin (10 μg), sulfamethoxazole/trimethoprim (SXT) (1.25/23.75 μg), and tetracycline (30 μg) (Hardy Diagnostics, Santa Maria, CA; BD Diagnostics, Sparks, MD). The zones of inhibition were measured, and isolates were classified as resistant or susceptible according to CLSI ranges; isolates classified as intermediate are also reported as ‘resistant’ in this study. Quality control checks were performed between every 25 tests using *E. coli* strain ATCC 25922. An isolate was considered multi-drug resistant (MDR) if resistant to three or more of the antibiotics tested.

Prevalence of AMR and MDR phenotypes is defined by the presence of resistance within any of the isolates collected from a single sample. The multi-antibiotic resistance (MAR) index of each sample was calculated as *a*/(*b*×*c*), where *a* is the total number of resistances observed within isolates collected from the sample, *b* is the number of the tested antibiotics (*n* = 8) and, *c* is the number of *E. coli* isolates collected from that sample (Krumperman, 1983).

### 2.5. Extended-spectrum beta-lactamase (ESBL) testing

*E. coli* isolates were tested for the production of ESBL enzyme using the combination disk test (CLSI, 2020). Zones of inhibition to disks containing cefotaxime (30 μg), ceftazidime (30 μg), cefotaxime/clavulanic acid (30/10 μg), and ceftazidime/clavulanic acid (30/10 μg) were measured (BD Diagnostics, Sparks, MD). A difference in zone of inhibition of ≥ 5 mm diameter between antibiotic and antibiotic-acid disk combinations signified the production of ESBL enzyme. An internal quality control and *E. coli* ATCC 25922 strain were used as ESBL-positive and ESBL-negative controls, respectively.

### 2.6. Statistical analysis

All statistical analysis was performed using R (version 4.0.3, The R Foundation, Vienna, Austria). A Kruskal-Wallis rank test was used to compare antibiotic-resistant *E. coli* in wastewater influent in different wastewater treatment utilities. Pearson’s chi-square test and generalized estimating equation models in R “geepack” package were used to investigate dynamics of resistance prevalence of different phenotypes in influent compared to secondary effluent, final effluent, and biosolids while accounting for temporal autocorrelation and cluster effects. Prevalence of different AMR phenotypes were set as the response variables and wastewater flow (i.e., influent, secondary effluent, final effluent, or biosolids) as the predictive variable. To test for the impact of seasonal variability on AMR *E. coli* in wastewater influent, generalized linear models (GLM) were developed using cluster-robust variance estimators with R “clubSandwich” package. In these tests, prevalence of different AMR phenotypes was set as the response variable and the predictor variables were seasons (winter or summer) and the interaction of seasons and region (Eastern Oregon or coastal/Willamette Valley). A similar procedure was taken by developing a linear model for determining the impact of seasonal variations on MAR index in wastewater influent. Pearson’s chi-squared tests were performed on pairwise phenotypes to analyze the co-occurrence of different resistances in *E. coli* isolates. Statistical significance was defined using an α = 0.05. The Bonferroni correction was used to correct the p-values for multiple tests.

## 3. Results and discussion

### 3.1. Physical and chemical characteristics of wastewater treatment utilities

The average wastewater physical-chemical characteristics for each wastewater treatment utility are reported in Table S2. Near-neutral pH was detected in wastewater influent, secondary effluent, and final effluent. The average pH of biosolids was 8.5 ± 0.4 (average ± standard error) and 8.7 ± 0.4 in winter and summer, respectively. When data from alkaline treated biosolids were excluded, the average pH was 8.0 ± 0.3 in winter and 8.0 ± 0.2 in summer. Total suspended solids, volatile suspended solids, ammonia, and conductivity in the influent were observed to be on average higher in summer compared to winter. These levels (solids, ammonia, and conductivity) generally decreased from wastewater influent to final effluent.

### 3.2. Dynamics of antibiotic-resistant *E. coli* in different flows in Oregon wastewater utilities

A total of 1143 *E. coli* isolates were collected from wastewater influent, secondary effluent, final effluent, and treated biosolids in winter and summer of 2019 and 2020 from the 17 participating wastewater treatment utilities across Oregon (Figure S1). Overall, 31.6% (*n* = 361) of all *E. coli* isolates harbored resistance to at least one of the eight tested antibiotics. Of these, 57.3% (*n* = 207) were resistant to ampicillin, followed by 43.8% (*n* = 158), 42.9% (*n* = 155), 20.5% (*n* = 74), 20.2% (*n* = 73), 5.5% (*n* = 20), and 4.2% (*n* = 15) to tetracycline, streptomycin, ciprofloxacin, SXT, cefotaxime, and ceftazidime, respectively (Figure 1a). Only one isolate harbored resistance to imipenem which was collected from a secondary effluent sample (Figure 1b). Among the antibiotic-resistant *E. coli* isolates, 27.2% (*n* = 100) carried MDR phenotypes. In wastewater influent, 28.0% of isolated *E. coli* harbored resistance to at least one of the eight tested antibiotics, ranging from 31.3% in winter to 25.3% in summer, with similar rates between geographical regions (27.5% in Eastern Oregon vs. 28.2% in coastal and valley region; Figure 1b-e). Resistance to ampicillin (14.6%), tetracycline (14.6%), and streptomycin (12.4%) were the most prevalent AMR phenotypes in wastewater influent (Figure 1a). Of the 102 antibiotic-resistant *E. coli* isolates from wastewater influent, 20.6% harbored MDR phenotype with similar rates between seasons or geographical regions (Figure 1b-e). In secondary effluent, we report that 31.4% of *E. coli* isolates harbored resistance to at least one antibiotic, of which 29.0% carried MDR phenotypes (Figure 1a). The AMR rates in secondary effluent were similar between winter and summer or between Eastern Oregon and the coastal/valley regions. Our data on *E. coli* isolates from final effluent shows 31.7% resistance to at least one of the eight tested antibiotics (Figure 1a). AMR rates in final effluent ranged from 42.1% in winter to 20.0% in summer with 26.7% in Eastern Oregon to 33.1% in the coastal and valley region (Figure 1b-e). Of the 65 *E. coli* isolates that harbored resistance to at least one of the tested antibiotics in final effluent, 27.7% (*n* = 18) were MDR. Similar to the influent flow, our findings highlight ampicillin (21.3%), streptomycin (14.4%), and tetracycline (11.9%) as the antibiotics least effective against *E. coli* isolated from final effluent. Finally, of the 259 *E. coli* colonies isolated from biosolids 36.2% were resistant to at least one of the eight tested antibiotics. We observed the highest rates of AMR in biosolids. Biosolids isolates of *E. coli* were resistant to ampicillin (19.7%), tetracycline (15.4%), ciprofloxacin (15.4%), streptomycin (13.9%). Moreover, 34.0% of antibiotic-resistant *E. coli* colonies collected from biosolids showed MDR phenotypes.

**Figure 1.**
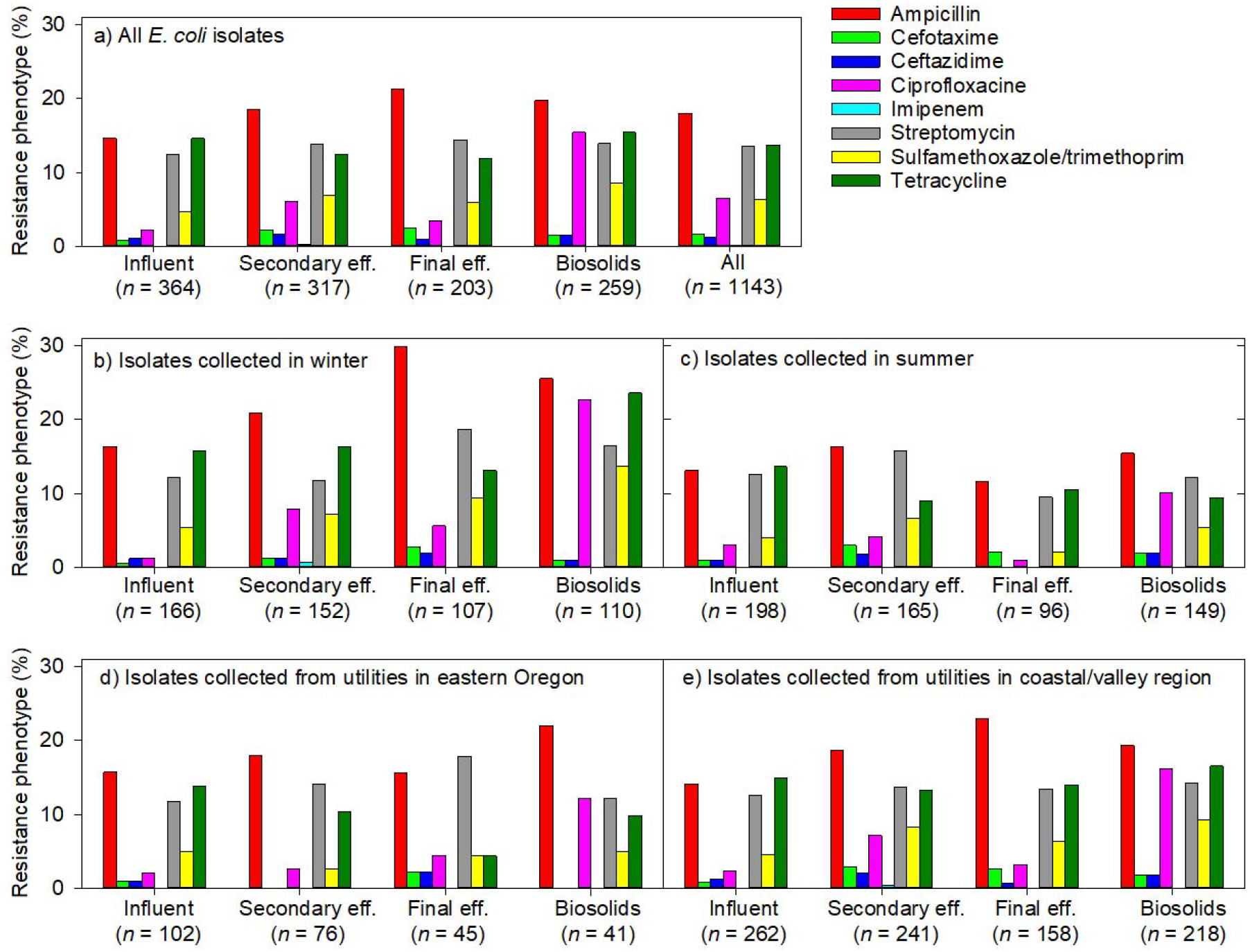
Percentage of antibiotic resistance phenotypes of *E. coli* isolates (*n* = 1143) collected from influent, secondary effluent, final effluent, and treated biosolids from wastewater treatment utilities across Oregon (*n* = 5 in Eastern Oregon, *n* = 12 in Willamette Valley/coastal regions) in winter and summer over two years (2019 and 2020). Resistance phenotype is displayed as percentage of isolates with intermediate to resistant classification to eight different antibiotic(s) based on CLSI guidelines (2020).

In collected samples (*n* = 246), AMR phenotypes were prevalent in 81.0% of influent samples (*n* = 63), 77.8% of secondary effluent (*n* = 63), 52.4% of final effluent (*n* = 63), and 71.9% of biosolids (*n* = 57) (Figure 2a). Considering dynamics of the AMR phenotypes’ pressure between wastewater flows (i.e., influent compared to secondary effluent, final effluent, and biosolids), higher prevalence of ciprofloxacin-resistant *E. coli* was observed in secondary effluent (28.6%; *p* = 0.02; *p* > 0.05 with Bonferroni correction; Table S3a-b) and biosolids (33.3%, *p* < 0.001; *p* < 0.01 with Bonferroni correction; Table S3a-b) compared to influent (12.7%) (Figure 2a, Table S3a and b). *E. coli* isolated from final effluent harbored significantly lower (20.6%, *p* = 0.004; *p* < 0.05 with Bonferroni correction; Table S3a-b) tetracycline resistance phenotype as compared to influent flow (52.4%). When excluding missing data (i.e., only including utilities B, D-J, M-Q with samples from summer 2019, winter 2020, and summer 2020; *n* = 153) results demonstrated higher prevalence of ciprofloxacin-resistant *E. coli* in biosolids compared to influent (*p* = 0.01; *p* > 0.05 with Bonferroni correction; Table S3a and b). Moreover, among all samples (*n* = 246), MAR index ranged between 0 to 0.38 with an average of 0.08 ± 0.01 (Figure 3). The average MAR index was 0.06 ± 0.01 in influent and 0.08 ± 0.01 in secondary effluent and final effluent. MAR index of 0.2 has been suggested as a threshold indicator for an environment with high-risk AMR exposure (Harnisz et al., 2011). In our study, of the 246 sampling incidences, 21 samples from 11 wastewater treatment utilities exhibited a MAR index above 0.2: two influent, six secondary effluent, seven final effluent, and six biosolids (Figure 3).

**Figure 2.**
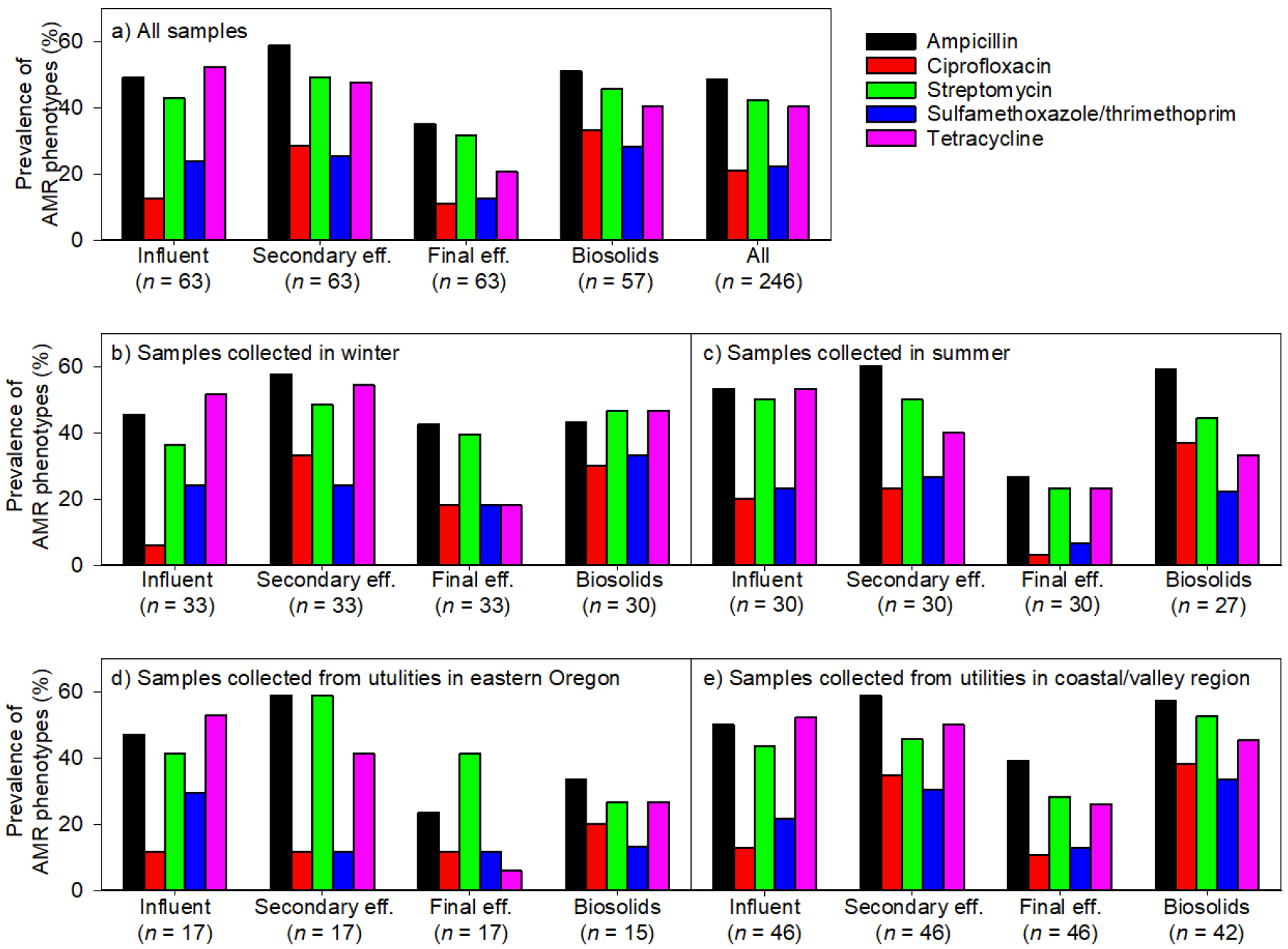
Prevalence of antibiotic-resistant *E. coli* in different flows (i.e., influent, secondary effluent, final effluent, and biosolids) from wastewater treatment utilities across Oregon (*n* = 5 in Eastern Oregon, *n* = 12 in Willamette Valley/coastal regions) in winter and summer over two years (2019 and 2020). Prevalence of resistance phenotype is displayed as presence/absence based on resistance within any of the isolates collected from a single sample. AMR: antimicrobial resistance (i.e., resistance to ≥1 of the tested antibiotics).

**Figure 3.**
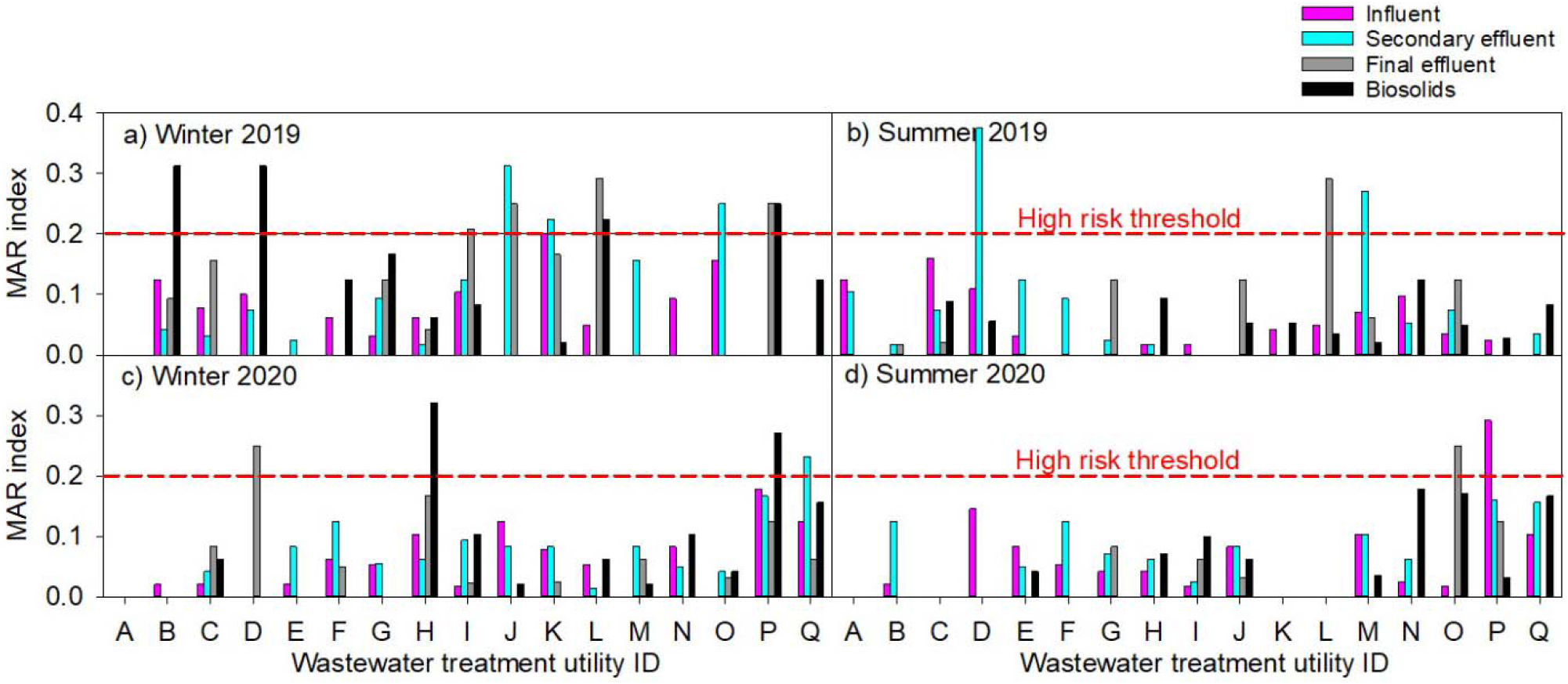
Multi-antibiotic resistance (MAR) index of *E. coli* isolated from wastewater influent, secondary effluent, final effluent and biosolids. Samples were collected from 17 wastewater treatment utilities across Oregon during winter and summer over two years (2019 and 2020).

### 3.3. Impact of seasonal variations on antibiotic-resistant *E. coli* in wastewater influent

We studied the impact of seasonal variations (winter vs summer) while controlling for regional effects (Eastern Oregon vs coastal/valley) on AMR *E. coli* in wastewater influent (*n* = 63). Our data shows significantly higher prevalence of ciprofloxacin-resistant *E. coli* in influent in summer samples as compared to winter (*p* < 0.001; *p* < 0.05 with Bonferroni correction; Table 1; Figure 2b-c). In addition, in winter season, the prevalence of ciprofloxacin resistance in influent collected from coastal/valley was higher (*p* < 0.001; *p* < 0.05 with Bonferroni correction; Table 1; Figure 2b-e) than Eastern Oregon samples (Table 1). To further investigate the impact of seasonal variations on antibiotic-resistant *E. coli* in influent, we focused on summer 2019 and winter 2020 samples, excluding seasons with missing samples (*n* = 34; Figure S1). When only comparing influent samples between summer 2019 and winter 2020, MAR index, ampicillin resistance prevalence, and ciprofloxacin resistance prevalence were significantly higher in summer compared to winter (*p* = 0.02, <0.001, and <0.001, respectively, with *p* < 0.05 with Bonferroni correction for all three tests; Table 1). When controlled for regional variations in winter influent samples, higher MAR index was observed in coastal/valley compared to Eastern Oregon over both seasons (*p* = 0.02 in winter and *p* = 0.03 in summer; *p* > 0.05 for both seasons with Bonferroni correction; Table 1; Figure 3). Controlled for regional variations, findings also demonstrate significantly higher ampicillin resistance prevalence, ciprofloxacin resistance prevalence, and SXT resistance prevalence in coastal/valley compared to Eastern Oregon (*p* < 0.001 and *p* < 0.05 with Bonferroni correction for the other three tests; Table 1; Figure 2b-e).

**Table 1.**
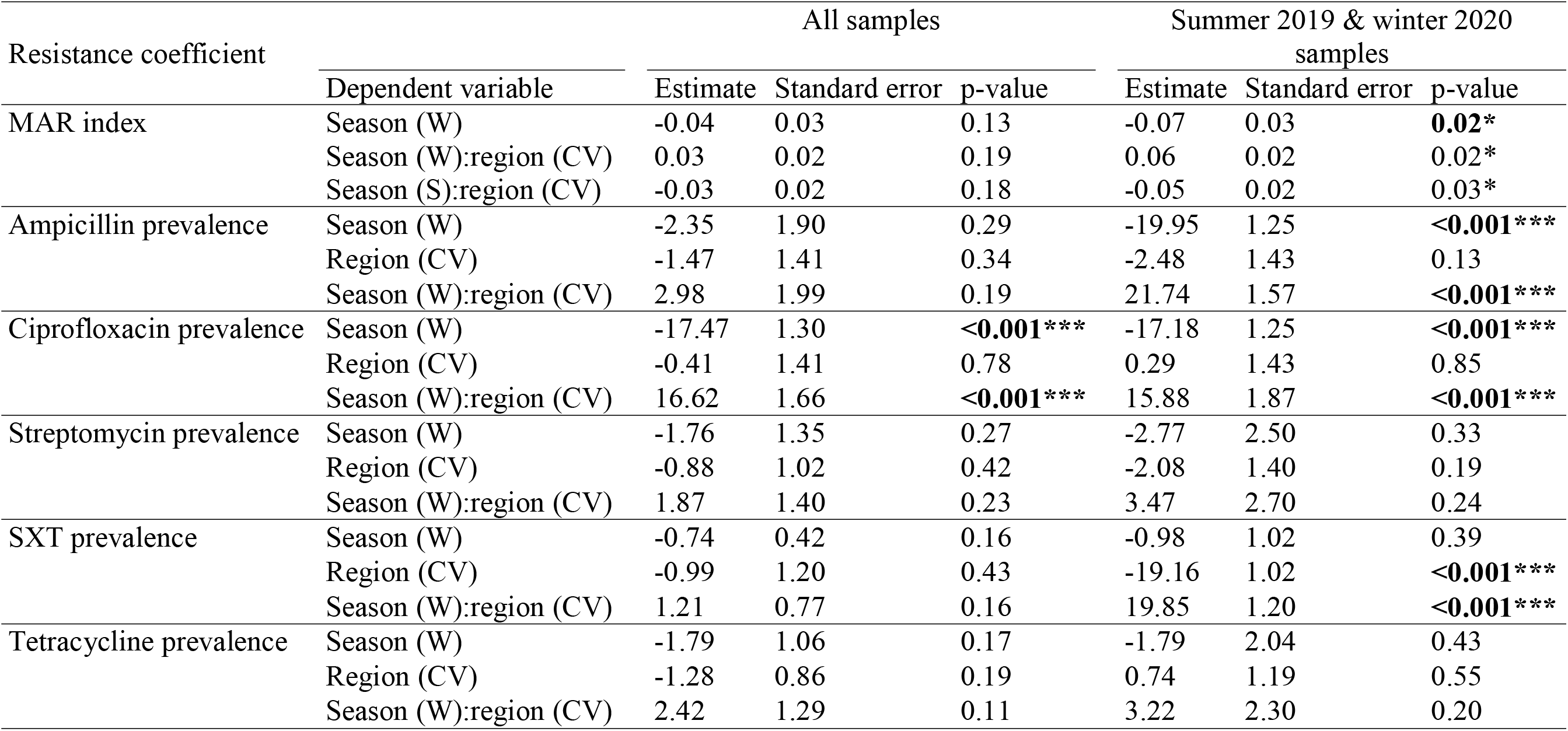
Seasonal variability of AMR *E. coli* in wastewater influent. Linear model’s estimates and standard errors reported for MAR index in wastewater influent. Generalized linear models and cluster-robust variance estimates and standard errors reported for prevalence of different antibiotic resistance phenotypes. Seasons (winter (W) and summer (S)) and interaction between seasons and region (Eastern Oregon (E) and coastal/valley (CV)) were set as predictor variables. Analyses are shown for all samples as well as samples collected in summer 2019 and winter 2020, excluding seasons with missing samples. SXT: sulfamethoxazole/trimethoprim. ^*^ *p* < 0.05, ^**^ *p* < 0.01, ^***^ *p* < 0.001. Significant correlations with Bonferroni correction are highlighted in bold.

Different observations on the seasonal variability of ciprofloxacin have been previously reported. For example, ciprofloxacin resistance was not impacted by seasonal changes in the blood stream isolates of *E. coli* (Ramsey et al., 2019), whereas higher rates were reported in winter in urinary tract infection studies (Soucy et al., 2020). Accordingly, there are higher prescriptions of ciprofloxacin antibiotic over summer associated with higher rates of urinary tract infections as well as skin and soft tissue infections (Durkin et al., 2018). Ciprofloxacin resistance has been increasing over the past 20 years and has been associated with higher usage of this antibiotic in inpatient care units and outpatient treatments in the U.S. (Sanchez et al., 2012).

### 3.4. Impact of treatment process on antibiotic-resistant *E. coli*

Calculated MAR index is shown for each flow and utility based on season and collection year (Figure 3). In both seasons, the averages for MAR index were slightly higher in the secondary effluent compared to the influent. We observed a higher mean MAR index in secondary effluent flow compared to the influent in eight of the 13 wastewater treatment utilities that used conventional biological treatment processes (i.e., activated sludge or lagoon). An increase in average MAR index from influent to secondary effluent was also observed in two of the four utilities with advanced treatment systems (i.e., membrane bioreactor and elevated nutrient removal). Our findings demonstrate increases of MAR index from influent to secondary effluent in 11 of the 17 wastewater treatment utilities, including those with advanced treatment systems, delineating the potential proliferation of AMR in biological treatment systems. This is contrary to several reports showing effective AMR removal during advanced biological treatments, including membrane bioreactors and advanced nutrient removal processes (Korzeniewska and Harnisz, 2018; Munir et al., 2011). Even though increases in antibiotic-resistant bacteria and determinant genes after biological treatment has been previously reported (Honda et al., 2020; Kumar et al., 2020; Mezrioui and Baleux, 1994; Zhang et al., 2009), the potential link between the role of the treatment process and proliferation of AMR remains unclear.

In all utilities, chlorination and/or UV were used for disinfection of the secondary effluent, after which 32.0% of the collected *E. coli* colonies were resistant to at least one of the eight tested antibiotics (27.2% of which were MDR) in the final effluent. Amongst the wastewater treatment utilities applying UV disinfection, the average MAR index did not change (∼0.12) from secondary effluent to final effluent. Comparatively, in wastewater treatment utilities utilizing chlorine, MAR index decreased slightly from secondary effluent (0.10) to final effluent (0.08).

In treated biosolids (collected after anaerobic digestion, aerobic treatment, alkaline treatment, or mechanical treatment), 36.9% of the isolates harbored AMR with highest rates of MDR in comparison to other flows’ phenotypes (34.0% of AMR population). The average MAR index in biosolids was 0.13 ± 0.02 in winter and 0.07 ± 0.01 in summer.

### 3.5. Co-occurrence of antibiotic-resistant *E. coli* phenotypes

While most (68.4%) of the 1143 *E. coli* isolates in our study were susceptible to all eight tested antibiotics, 31.6% of the *E. coli* isolates (*n* = 361) exhibited resistance to at least one antibiotic. Of these, 27.7% (*n* = 100) carried MDR phenotypes. In influent, 28.0% (*n* = 102) of *E. coli* isolates were resistant to at least one antibiotic and 5.8% (*n* = 21) showed MDR phenotypes (Figure 4a). In secondary effluent, 31.6% (*n* = 100) of isolated *E. coli* harbored resistance against tested antibiotics and 9.2% (*n* = 29) were MDR (Figure 4a). In final effluent and biosolids, 32.0% (*n* = 65) and 36.3% (*n* = 94) of *E. coli* isolates were categorized as AMR and 8.9% (*n* = 18) and 12.4% (*n* = 32) were MDR, respectively (Figure 4a). Among the 100 MDR *E. coli* isolates, 59.0% were resistant to three antibiotics, whereas 27.0%, 8.0%, and 6.0% of the MDR isolates harbored resistances to four, five, and six antibiotics, respectively.

**Figure 4.**
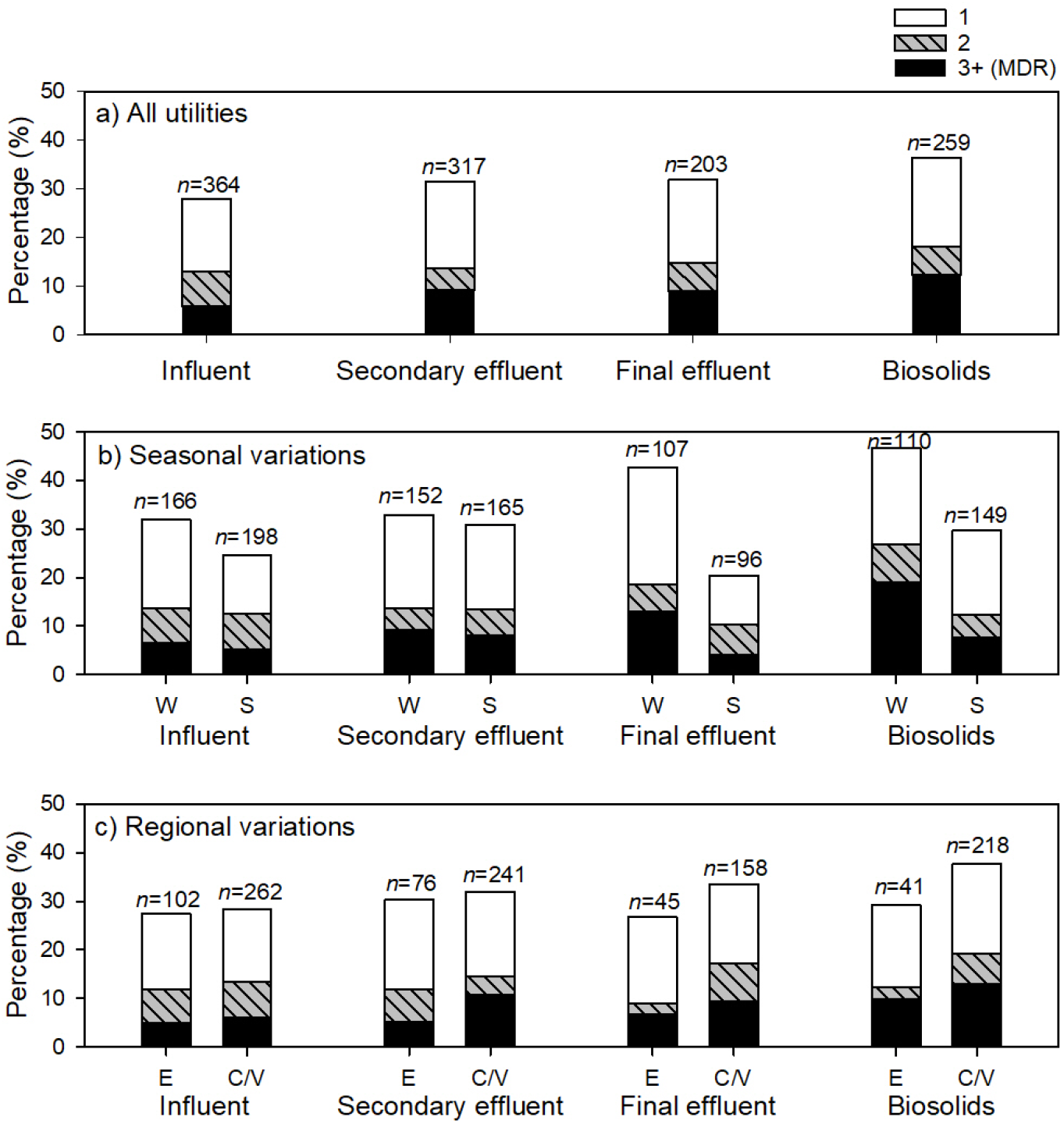
Percentage of *E. coli* isolates resistant to 1, 2, and 3 or more (multi-drug resistance; MDR) classes of antibiotics. *E. coli* isolated were collected from influent, secondary effluent, final effluent, and biosolids for a) all utilities (*n* = 17), b) samples collected in winter (W) and summer (S), and c) samples collected from wastewater treatment utilities in Eastern Oregon (E) and coastal/valley (C/V) regions of Oregon. The total number of samples are shown above each bar.

Resistance to ampicillin was the most common phenotype among MDR isolates (observed in 91% of MDR isolates); followed by resistances to tetracycline (82.0%), streptomycin (72.0%), and SXT (64.0%). The most common MDR phenotype combination included simultaneous resistances to ampicillin, streptomycin, and tetracycline (*n* = 49), followed by ampicillin, streptomycin, and SXT (*n* = 46).

Co-occurrences of resistance to tested antibiotics were assessed between the *E. coli* colonies that harbored at least two AMR phenotypes (*n* = 169). Resistance to cefotaxime, ceftazidime, and imipenem were observed in only 20, 15, and 1 *E. coli* isolates, respectively, and were excluded from the analysis of AMR phenotypes’ co-occurrences because of low statistical power. The occurrence of SXT-resistant *E. coli* showed significant positive correlations with ampicillin, streptomycin, and tetracycline (*p* < 0.001 with Bonferroni correction; Table 2).

**Table 2.** Co-occurrences of different phenotypes of antibiotic resistance in wastewater treatment utilities. Chi-square statistic values (p-values) are reported for chi-square tests for all isolates collected from different wastewater flows (i.e., influent, secondary, effluent, and biosolids). SXT: sulfamethoxazole/trimethoprim. ^*^ *p* < 0.05, ^**^ *p* < 0.01, ^***^ *p* < 0.001. Significant correlations with Bonferroni correction are highlighted in bold.

| Resistance phenotypes | Ampicillin | Ciprofloxacin | Streptomycin | SXT |
| --- | --- | --- | --- | --- |
| Ampicillin |  |  |  |  |
| Ciprofloxacin | 1.07 (0.03*) |  |  |  |
| Streptomycin | 4.98 (0.03*) | 4.75 (0.03*) |  |  |
| SXT | 19.44<br>(**<0.001\*\*\***) | 2.17 (0.14) | 23.10<br>(**<0.001\*\*\***) |  |
| Tetracycline | 0 (1.0) | 0.67 (0.41) | 0.62 (0.43) | 32.4<br>(**<0.001\*\*\***) |

Moreover, of the 73 *E. coli* isolates with resistance to SXT, 87.7% (*n* = 64) harbored MDR phenotype. Prevalence of SXT-resistance in *E. coli* has been previously associated with occurrence of class 1 integron genes, indicator of horizontal gene transfer (Shin et al., 2015). Moreover, co-occurrences of resistance were significant between ampicillin and ciprofloxacin and with streptomycin (*p* = 0.03 for all three correlations; *p* > 0.05 with Bonferroni correction; Table 2).

### 3.6. ESBL-producing *E. coli* in wastewater treatment utilities across Oregon

Our study is the first to report on ESBL-producing *E. coli* isolates in Oregon wastewater systems and the first to report this data for biosolids from the U.S. Of the 1143 *E. coli* colonies isolated from 246 samples, 1.2% tested positive for ESBL-producing enzyme (*n* = 13). These isolates were collected from nine different wastewater treatment utilities. The distribution of ESBL-producing isolates was fairly even between influent (*n* = 3), secondary effluent (*n* = 2), final effluent (*n* = 4), and biosolids (*n* = 4). Among the ESBL-producing *E. coli*, six were detected in winter and seven in summer. Of these 13 isolates, 11 were collected from the Oregon coast and the Willamette Valley, of which eight belonged to wastewater utilities with higher serving population densities. Moreover, 12 of the 13 ESBL-producing *E. coli* isolates harbored MDR phenotypes, of which seven showed resistance to six or more antibiotics. Among these ESBL-producing isolates, cefotaxime resistance was the most common AMR phenotype (*n* = 13) followed by ampicillin (*n* = 11), tetracycline (*n* = 11), ceftazidime (*n* = 10), ciprofloxacin (*n* = 9), SXT (*n* = 9), and streptomycin (*n* = 8). All ESBL-producing *E. coli* isolates were susceptible to imipenem. The only other study in the U.S., to our knowledge, on ESBL-producing *E. coli* in wastewater systems (primary clarifier and final effluent) reported 20% prevalence of carbapenemase, ESBL-associated genes, or both in antibiotic-resistant *E. coli* isolates collected in the states of New Jersey, Maryland, Ohio, Texas, Colorado, California (Hoelle et al., 2019). It is also important to note that we detected only one imipenem-resistant *E. coli* isolate, collected from a secondary effluent in winter. Carbapenems, such as imipenem, are the common treatment option for infections with ESBL-producing bacteria. Hence, the occurrence of imipenem-resistant bacteria in the environment suggests clinical resistance to last resort antibiotics. ESBL-producing bacterial infections have been previously correlated with hospital stays of over six times longer than infections with non-ESBL-producing bacteria (Lautenbach et al., 2001).

Longer hospital stays and delayed diagnoses invite more opportunity for lasting damage from these infections.

## 4. Conclusion

This study sheds light on the abundance and diversity of antibiotic-resistant *E. coli* in 17 wastewater treatment utilities across Oregon over winter and summer seasons. Amongst the 1143 collected *E. coli* isolates from 246 samples, AMR phenotypes were detected in *31*.*6*% of the *E. coli* isolates from different flows and seasons throughout the study, with some isolates showing resistance to up to six classes of antibiotics. MDR phenotypes were observed among 27.7% of isolates with AMR phenotypes, and the most common combinations included simultaneous resistances to ampicillin, streptomycin, and tetracycline (*n* = 49), followed by ampicillin, streptomycin, and SXT (*n* = 46). Among tested phenotypes, ampicillin resistance was the most common AMR phenotype, followed by streptomycin and tetracycline. Significant correlations were observed between resistance to SXT and resistances to ampicillin, ciprofloxacin, and tetracycline (*p* < 0.001). Among collected *E. coli* isolates, 13 ESBL-positive and one imipenem resistance were detected. When analyzing the resistance phenotypes between different flows (i.e., influent, secondary effluent, final effluent, and biosolids), ciprofloxacin resistance was significantly more prevalent in biosolids compared to influent (*p* <0.05) and tetracycline resistance was significantly lower in effluent as compared to influent (*p* <0.05). Seasonal impact on AMR *E. coli* in wastewater influent was observed through significantly higher MAR index, ampicillin resistance prevalence, and ciprofloxacin resistance prevalence in summer compared to winter (*p* < 0.05). This state-wide study confirms the widespread proliferation of antibiotic-resistant, MDR, and ESBL-producing *E. coli* and further identifies wastewater surveillance as an epidemiological screening and identification tool.

## Data Availability

All data produced in the present work are contained in the manuscript or can be available upon reasonable request to the corresponding author.

## Declaration of Competing Interest

The authors declare that they have no known competing financial interests or personal relationships that could have appeared to influence the work reported in this paper.

## Acknowledgement

This work was supported by the USDA National Institute of Food and Agriculture, Agricultural and Food Research Initiative Competitive Program, Agriculture Economics and Rural Communities, Grant No. 2018-67017-27631.

## Notes

### Competing Interest Statement

The authors have declared no competing interest.

