## supplementary material for "Antibiotic resistance profile of *E. coli* isolates in 17 municipal wastewater utilities across Oregon"

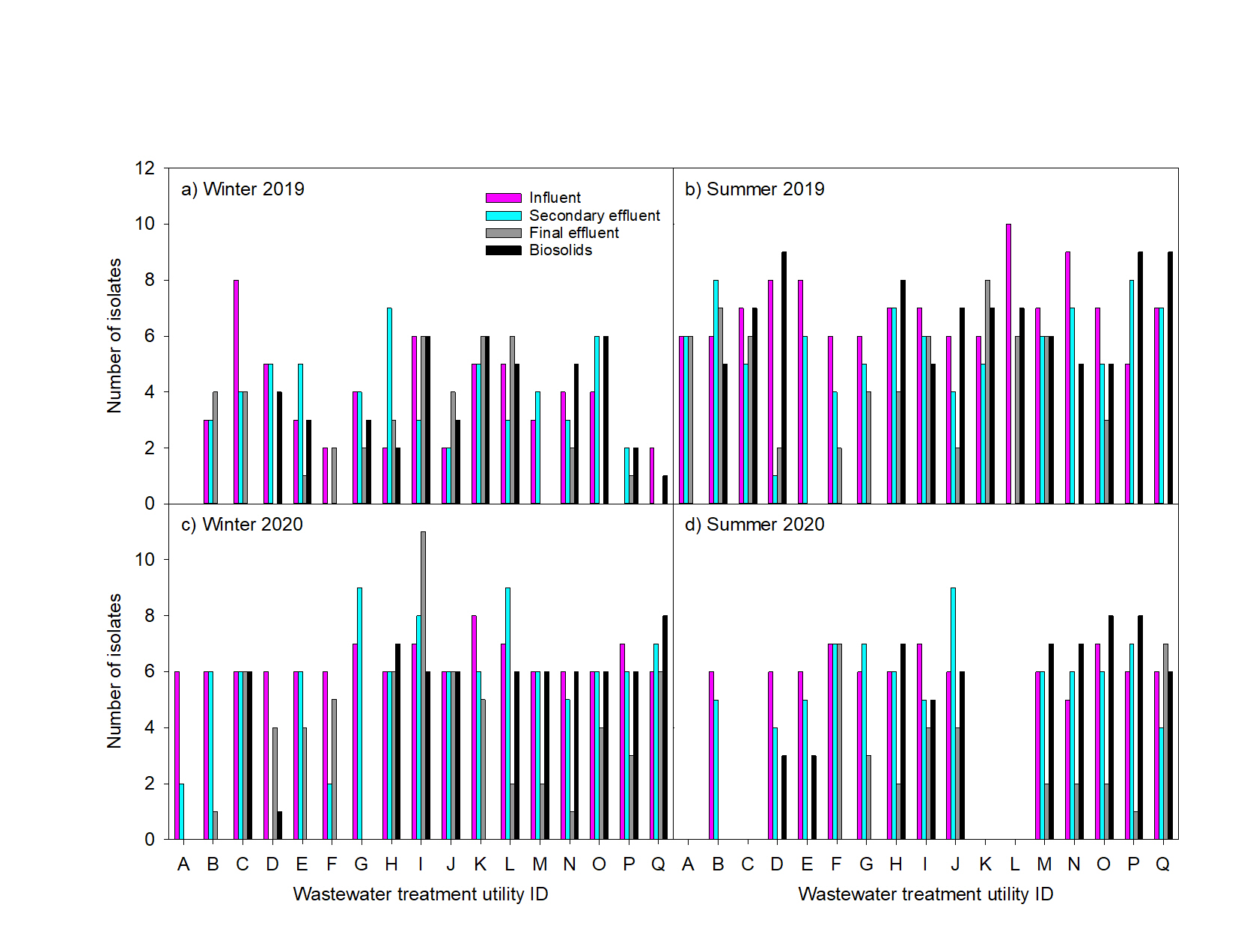

Figure S1. Number of *E. coli* isolates collected from different flows (influent, secondary effluent, final effluent, and biosolids) of 17 wastewater treatment utilities across Oregon during winter and summer over two years (2019 and 2020). No biosolids were collected from utility F in summer 2019. No samples were collected from utility A in winter 2019, nor from utilities A, C, F, and K in summer 2020. No *E. coli* colonies were observed for collection in other samples with zero number of isolates.

Table S1. Oregon wastewater treatment utilities (*n* = 17) that participated in sample collection to characterize antibiotic-resistant *Escherichia coli* in various flows.

| Facility | Region^a^ | Population Served^b^ | Biological Treatment^c^ | Disinfection^d^ | Sludge Treatment^e^ |
| --- | --- | --- | --- | --- | --- |
| A | E | S | Conv | Cl | AD |
| B | E | S | Conv | Cl | Alk |
| C | E | S | Adv | Cl | AD |
| D | E | S | Adv | Cl | AD |
| E | E | S | Adv | Cl | AD |
| F | V | S | Conv | Cl | AD |
| G | C | S | Conv | Cl | Alk |
| H | V | S | Conv | Cl | Mech |
| I | V | S | Conv | UV | AD |
| J | V | S | Conv | UV | Oxic |
| K | C | S | Conv | UV | Oxic |
| L | V | L | Conv | Cl | AD |
| M | V | L | Conv | Cl | AD |
| N | V | L | Conv | Cl | AD |
| O | V | L | Conv | Cl/UV | AD |
| P | V | L | Adv | Cl | AD |
| Q | V | L | Adv | Cl | AD |

^a^Region: C – coastal, E – eastern Oregon, V – Willamette Valley.

^b^Population served: L – large (80,000-700,000), S – small (2,000-80,000).

^c^Biological treatment: Adv – advanced (biological nutrient removal, membrane bioreactor), Conv – conventional (lagoon, activated sludge, oxidation ditch).

^d^Disinfection: Cl – chlorine, UV – ultraviolet.

^e^Sludge Treatment: AD – anaerobic digestion, Alk – alkaline, Mech – mechanical dewatering, Oxic – aerobic.

Table S2. Physical-chemical characteristics (mean ± standard deviation) of wastewater influent (I), secondary effluent (S), final effluent (E), and biosolids (B) collected from wastewater treatment utilities in Oregon (n = 17). Values are average ± standard error for samples (n = 3-4) collected over two seasons (winter vs. summer) in 2019 and 2020.

| Utility ID | Flow | n | pH | Conductivity  (µS/cm) | TSS  (mg/L) | VSS  (mg/l) | Total Solids (%) | Ammonia (mg/L) |
| --- | --- | --- | --- | --- | --- | --- | --- | --- |
| A | I | 3 | 7.7 ± 2.2 | 559.7 ± 215.2 | 120.9 ± 62.8 | 91.6 ± 64.3 | — | 14.8 ± 7.6 |
|  | S | 3 | 8.8 ± 2.6 | 414.3 ± 151.2 | — | — | — | — |
|  | E | 3 | 8.4 ± 2.4 | 567.3 ± 250.5 | 34.9 ± 18.1 | 33.6 ± 16.9 | — | 1.6 ± 1.3 |
|  | B | 0 | — | — | — | — | — | — |
| B | I | 4 | 8.0 ± 1.8 | 801.8 ± 190.3 | 245.3 ± 22.7 | 225.8 ± 23.5 | — | 46.1 ± 11.6 |
|  | S | 4 | 7.6 ± 1.7 | 403.5 ± 95.1 | — | — | — | — |
|  | E | 4 | 8.0 ± 1.8 | 1542.8 ± 956.1 | 5.2 ± 2.5 | 6.0 ± 1.9 | — | 0.0 0.3 |
|  | B | 4 | 13.1 ± 2.9 | 5820.0 ± 1518.4 | — | — | 85.8 ± 0.8 | — |
| C | I | 3 | 7.6 ± 2.2 | 1077.0 ± 312.1 | 268.3 ± 40.9 | 193.7 ± 35.8 | — | 30.9 ± 16.4 |
|  | S | 3 | 7.3 ± 2.1 | 676.3 ± 228.5 | — | — | — | — |
|  | E | 3 | 7.8 ± 2.3 | 750.7 ± 274.2 | 867.1 ± 859.1 | 868.7 ± 860.7 | — | 0.0 ± 0.1 |
|  | B | 3 | 8.7 ± 2.5 | 711.7 ± 241.5 | — | — | 75.3 ± 6.1 | — |
| D | I | 4 | 8.0 ± 1.8 | 1588.5 ± 423.3 | 172.9 ± 23.2 | 158.5 ± 24.8 | — | 40.4 ± 9.6 |
|  | S | 4 | 8.2 ± 1.8 | 979.0 ± 279.9 | — | — | — | — |
|  | E | 4 | 8.0 ± 1.8 | 990.0 ± 236.5 | 14.9 ± 7.5 | 11.3 ± 8.9 | — | 0.0 ± 0.2 |
|  | B | 4 | 8.7 ± 2.0 | 857.0 ± 259.4 | — | — | 85.5 ± 1.0 | — |
| E | I | 4 | 7.9 ± 1.8 | 831.0 ± 199.3 | 154.0 ± 29.4 | 372.5 ± 105.8 | — | 35.1 ± 7.9 |
|  | S | 4 | 7.9 ± 1.8 | 654.5 ± 152.2 | — | — | — | — |
|  | E | 4 | 7.9 ± 1.8 | 701.8 ± 186.6 | 21.5 ± 4.4 | 18.3 ± 3.6 | — | 9.7 ± 5.6 |
|  | B | 4 | 7.0 ± 1.6 | 1417.5 ± 535.9 | — | — | 46.3 ± 17.3 | — |
| F | I | 4 | 7.9 ± 1.8 | 799.8 ± 225.7 | 144.2 ± 23.0 | 146.3 ± 32.7 | — | 27.2 ± 8.5 |
|  | S | 4 | 7.6 ± 1.7 | 689.5 ± 214.9 | — | — | — | — |
|  | E | 4 | 7.6 ± 1.7 | 520.8 ± 119.8 | 51.2 ± 34.4 | 41.4 ± 24.7 | — | 14.5 ± 7.9 |
|  | B | 0 | — | — | — | — | — | — |
| G | I | 4 | 7.4 ± 1.6 | 776.3 ± 226.3 | 172.3 ± 20.6 | 155.9 ± 20.1 | — | 21.8 ± 5.6 |
|  | S | 4 | 7.5 ± 1.7 | 637.5 ± 152.8 | — | — | — | — |
|  | E | 4 | 7.7 ± 1.7 | 571.5 ± 154.9 | 1.5 ± 1.0 | 3.0 ± 1.0 | — | 9.7 ± 3.9 |
|  | B | 4 | 11.4 ± 2.8 | 6500.0 ± 1807.0 | — | — | 77.5 ± 2.3 | — |
| H | I | 4 | 7.4 ± 1.7 | 731.3 ± 175.9 | 335.5 ± 188.5 | 292.3 ± 161.1 | — | 31.4 ± 7.8 |
|  | S | 4 | 7.3 ± 1.6 | 703.8 ± 253.5 | — | — | — | — |
|  | E | 4 | 7.7 ± 1.7 | 1279.5 ± 763.1 | 3.0 ± 1.9 | 3.0 ± 0.7 | — | 7.0 ± 2.1 |
|  | B | 4 | 6.6 ± 1.5 | 722.2 ± 258.4 | — | — | 89.1 ± 3.3 | — |
| I | I | 4 | 7.5 ± 1.7 | 668.5 ± 175.9 | 350.7 ± 79.3 | 372.8 ± 105.8 | — | 35.1 ± 7.9 |
|  | S | 4 | 7.8 ± 1.8 | 456.4 ± 127.7 | — | — | — | — |
|  | E | 4 | 7.9 ± 1.8 | 414.5 ± 128.2 | 5.0 ± 2.3 | 5.0 ± 1.4 | — | 9.7 ± 5.6 |
|  | B | 4 | 8.6 ± 1.9 | 957.8 ± 237.2 | — | — | 67.4 ± 22.1 | — |
|  |  |  |  |  |  |  | (continued) | |
| Utility ID | Flow | n | pH | Conductivity  (µS/cm) | TSS  (mg/L) | VSS  (mg/l) | Total Solids (%) | Ammonia (mg/L) |
| J | I | 4 | 7.7 ± 1.7 | 845.3.7 ± 229.9 | 178.5 ± 26.1 | 193.5 ± 37.3 | — | 41.7 ± 10.3 |
|  | S | 4 | 7.6 ± 1.7 | 553.0 ± 178.3 | — | — | — | — |
|  | E | 4 | 7.8 ± 1.8 | 642.3 ± 154.9 | 4.0 ± 2.1 | 2.2 ± 3.4 | — | 9.2 ± 4.1 |
|  | B | 4 | 7.0 ± 1.7 | 1741.9 ± 867.6 | — | — | 96.9 ± 1.4 | — |
| K | I | 3 | 7.6 ± 2.2 | 489.7 ± 180.6 | 145.3 ± 44.7 | 133.9 ± 39.4 | — | 22.4 ± 7.0 |
|  | S | 3 | 7.5 ± 2.2 | 629.0 ± 198.8 | — | — | — | — |
|  | E | 3 | 7.7 ± 2.2 | 535.7 ± 158.6 | 6.0 ± 3.2 | 7.6 ± 1.5 | — | 13.8 ± 4.2 |
|  | B | 3 | 6.1 ± 2.2 | 1342.3 ± 861.7 | — | — | 122.7 ± 23.9 | — |
| L | I | 3 | 7.5 ± 2.2 | 502.3 ± 194.3 | 207.6 ± 15.9 | 188.0 ± 11.3 | — | 37.4 ± 10.8 |
|  | S | 3 | 8.0 ± 2.3 | 882.0 ± 270.8 | — | — | — | — |
|  | E | 3 | 7.9 ± 2.3 | 679.3 ± 243.6 | 13.8 ± 3.2 | 12.1 ± 1.6 | — | 35.6 ± 10.5 |
|  | B | 3 | 8.7 ± 2.5 | 816.3 ± 242.3 | — | — | 83.2 ± 1.6 | — |
| M | I | 4 | 7.6 ± 1.7 | 813.3 ± 223.7 | 183.4 ± 15.0 | 141.6 ± 41.6 | — | 41.6 ± 10.7 |
|  | S | 4 | 7.8 ± 1.8 | 901.8 ± 301.9 | — | — | — | — |
|  | E | 4 | 7.8 ± 1.7 | 594.5 ± 189.5 | 3.8 ± 1.4 | 4.3 ± 0.4 | — | 27.1 ± 7.0 |
|  | B | 3 | 8.5 ± 2.4 | 1252.0 ± 421.0 | — | — | 86.2 ± 0.4 | — |
| N | I | 4 | 7.6 ± 1.7 | 810.5 ± 214.1 | 264.5 ± 46.0 | 248.7 ± 42.8 | — | 40.3 ± 11.1 |
|  | S | 4 | 7.9 ± 1.8 | 653.8 ± 157.7 | — | — | — | — |
|  | E | 4 | 7.9 ± 1.8 | 614.5 ± 158.2 | 3.2 ± 2.1 | 1.9 ± 2.2 | — | 12.9 ± 3.6 |
|  | B | 4 | 7.4 ± 1.9 | 1480.3 ± 469.5 | — | — | 81.7 ± 4.0 | — |
| O | I | 4 | 8.1 ± 1.8 | 855.5 ± 237.1 | 225.1 ± 30.8 | 205.2 ± 30.3 | — | 42.8 ± 10.1 |
|  | S | 4 | 7.9 ± 1.8 | 669.3 ± 158.1 | — | — | — | — |
|  | E | 4 | 8.1 ± 1.8 | 707.3 ± 215.2 | 6.8 ± 0.7 | 6.2 ± 2.0 | — | 25.3 ± 5.8 |
|  | B | 4 | 8.8 ± 2.0 | 1092.3 ± 317.1 | — | — | 80.9 ± 2.1 | — |
| P | I | 4 | 7.6 ± 1.7 | 699.5 ± 195.5 | 199.5 ± 30.1 | 174.1 ± 28.3 | — | 36.6 ± 10.8 |
|  | S | 4 | 7.4 ± 1.7 | 515.5 ± 136.2 | — | — | — | — |
|  | E | 4 | 7.6 ± 1.7 | 563.0 ± 150.5 | 1.3 ± 0.6 | 2.3 ± 0.4 | — | 1.9 ± 1.2 |
|  | B | 4 | 8.6 ± 1.9 | 1105.8 ± 311.0 | — | — | 82.6 ± 4.0 | — |
| Q | I | 4 | 7.8 ± 1.7 | 1283.3 ± 442.7 | 239.8 ± 84.1 | 194.9 ± 59.7 | — | 27.5 ± 9.0 |
|  | S | 4 | 7.3 ± 1.6 | 863.8 ± 233.7 | — | — | — | — |
|  | E | 4 | 7.7 ± 1.7 | 941.8 ± 293.4 | 1.3 ± 1.0 | 1.7 ± 1.5 | — | 3.1 ± 1.8 |
|  | B | 4 | 8.7 ± 1.9 | 798.3 ± 215.6 | — | — | 77.3 ± 0.4 | — |

Supplementary Table 3a. AMR prevalence in wastewater treatment utilities, correlations between different flows (i.e., influent, secondary, effluent, biosolids) shown for all utilities and for utilities with complete sample sets (i.e., utilities B, D-J, M-Q in S19, W20, S20, excluding utilities with missing samples). Chi-square statistic values and p-values are reported for chi-square tests for all utilities and for samples collected from utilities with complete sample sets B, D-J, M-Q in summer 2019 (S19), winter 2020 (W20), and summer 2020 S20), excluding seasons and utilities with missing samples. SXT: sulfamethoxazole/trimethoprim. * *p* < 0.05, ** *p* < 0.01, *** *p* < 0.001.

| Resistance phenotype | All utilities | |  | Utilities with complete sample sets | |
| --- | --- | --- | --- | --- | --- |
|  | X^2^ | p-value |  | X^2^ | p-value |
| Ampicillin | 5.00 | 0.17 |  | 5.51 | 0.14 |
| Ciprofloxacin | 15.36 | 0.002** |  | 8.97 | 0.03* |
| Streptomycin | 2.14 | 0.54 |  | 3.28 | 0.35 |
| SXT | 3.35 | 0.34 |  | 5.64 | 0.13 |
| Tetracycline | 8.84 | 0.03* |  | 5.13 | 0.16 |

Supplementary Table 3b. AMR *E. coli* prevalence in wastewater utilities, cluster effects compared to influent flow for tests with significant correlations in Supplementary Table 3a. Generalized estimating equation models’ estimates and standard errors reported for cluster effects comparison of secondary, effluent, and biosolids compared to influent flow. * *p* < 0.05, ** *p* < 0.01, *** *p* < 0.001. Significant correlations with Bonferroni correction are highlighted in bold.

| Resistance phenotype | Flow | Estimate | Standard error | p-value |
| --- | --- | --- | --- | --- |
| Ciprofloxacin resistance prevalence in all utilities | Secondary | 1.09 | 0.45 | 0.02* |
|  | Effluent | 0.13 | 0.49 | 0.79 |
|  | Biosolids | 1.67 | 0.45 | **<0.001***** |
| Tetracycline resistance prevalence in all utilities | Secondary | -0.09 | 0.29 | 0.76 |
|  | Effluent | -1.11 | 0.39 | **0.004**** |
|  | Biosolids | -0.19 | 0.39 | 0.63 |
| Ciprofloxacin resistance prevalence in utilities B, D-J, M-Q in summer 2019, winter 2020, and summer 2020 | Secondary | 0.81 | 0.57 | 0.15 |
|  | Effluent | 0.13 | 0.62 | 0.83 |
|  | Biosolids | 1.50 | 0.61 | 0.01* |
